# A 1 to 1000 SARS-CoV-2 reinfection proportion in members of a large healthcare provider in Israel: a preliminary report

**DOI:** 10.1101/2021.03.06.21253051

**Authors:** Galit Perez, Tamar Banon, Sivan Gazit, Shay Ben Moshe, Joshua Wortsman, Daniel Grupel, Asaf Peretz, Amir Ben Tov, Gabriel Chodick, Miri Mizrahi-Reuveni, Tal Patalon

## Abstract

With more than 100 million confirmed COVID-19 cases as of March 2021, reinfection is still considered to be rare. In light of increasing reports of reinfected COVID-19 patients, the need to better understand the real risk for reinfection is critical, with potential effects on public health policies aimed at containing the spread of SARS-CoV-2. In this descriptive preliminary report, we conducted a large-scale assessment on the country level of the possible occurrence of COVID-19 reinfection within the members of a large healthcare provider in Israel. Out of 149,735 individuals with a documented positive PCR test between March 2020 and January 2021, 154 had two positive PCR tests at least 100 days apart, reflecting a reinfection proportion of 1 per 1000. Given our strict inclusion criteria, we believe these numbers represent true reinfection incidence in MHS and should be clinically regarded as such.

## Introduction

In the ongoing effort to understand and control the coronavirus disease 2019 (COVID-19) global pandemic, one of the key questions that should be explored is how effectively the immune response protects a host from being re-infected with the Severe Acute Respiratory Syndrome Coronavirus 2 (SARS-CoV-2)^[1]^. The answer to this question has many implications including understanding if there is a real possibility to obtain herd immunity as well as significantly influencing policy-making in COVID-19 afflicted regions. This is particularly true when considering policy pertaining to social distancing practices and mask-wearing by individuals who were infected and recovered from SARS-CoV-2 ^[2]^. The widespread assumption that recovered patients develop a protective immune response has heavily influenced both research and policy-making, however, recent documentation of re-infected COVID-19 patients could potentially affect public health policies aimed at containing the spread of SARS-CoV-2.

With more than 100 million confirmed COVID-19 cases as of March 2021, reinfection is still considered to be a rare event^[3, 4]^. In light of increasing reports of reinfected COVID-19 patients, the need to better understand the real risk for reinfection is critical, with potential effects on public health policies aimed at containing the spread of SARS-CoV-2.

With the earliest cases of reinfection emerging in June 2020^[3]^, there is increasing documentation of patients who have been re-infected with SARS-CoV-2^[5]^. Re-infection has been determined either by genetic sequencing^[1, 5]^, which verifies that patients have been infected with two genetically distinct isolates of COVID-19, or by real-time polymerase chain reaction (PCR), demonstrating a newly positive PCR test months following recovery from COVID-19^[5, 6]^.

Several studies have indicated that re-infected patients typically experience milder symptoms during their second infection, possibly explained by priming of their adaptive immune response^[7, 8]^. However, case reports of individuals who have suffered a significantly more severe disease course during re-infection have also been documented^[5]^.

Admittedly, it is not surprising that there would be re-infections with SARS-CoV-2, as this phenomenon is known to occur with other coronaviruses^[6]^. Various strains of coronaviruses are responsible for the common cold and research displayed that the immune response to these viruses tend to develop quickly and wane over time which allows for re-infection to take place^[2]^. Although reinfections typically occur within one to three years with other coronaviruses, the increased re-exposure rate to SARS-CoV-2 may decrease the impact of the protective immune response and significantly shorten the time required for re-infection^[2, 6]^.

Within the Israeli population, at least one re-infection case has been documented, in August 2020^[9]^. Further research is therefore required to determine whether this case is an outlier or rather represents a trend of possible SARS-CoV-2 re-infection in the Israel population.

This descriptive preliminary report aims to perform a large-scale assessment on the country level of the possible occurrence of COVID-19 reinfection within the members of a large healthcare provider in Israel.

## Methods

### Data Source

This study was conducted using data from the Maccabi Healthcare Services (MHS) central computerized database, the second largest state-mandated not-for-profit healthcare provider in Israel, containing more than 2.5 million members (approximately 25% of the population) and is a representative sample of the Israeli population. This fully computerized database captures all information on patient interaction (including demographics, visits, diagnoses, procedures and more).

### Study population and design

In this retrospective cohort study, individuals were included if they had two positive PCR results, at least 100 days apart, from March 16, 2020 until January 27, 2021. The 100 days marker was selected in order to reduce the chance of prolonged shedding. The study was approved by the MHS institutional review board.

### Variables and definitions

The cohort was described by age (mean and standard deviation [±SD]), sex, socioeconomic status (SES), Immunocompromised registry (i.e. included in the MHS registry or not), the days between PCR tests (mean [±SD]), and COVID-19 related symptoms (±10 days from PCR test result), hospitalization and mortality. COVID-19 related symptoms included fever, cough, breathing difficulties, diarrhea, changes in the senses of taste or smell, fatigue, sore throat and headache.

Individuals’ ages were calculated at first positive PCR test date. SES of members’ residential area was based on a score ranked with 1 (lowest) to 10 built for commercial purposes by Points Location Intelligence using geographic information systems and data such as expenditures related to retail chains, credit cards and housing. This score is highly correlated with SES measured by the Israel Central Bureau of Statistics[10]. SES was categorized into low (1-4), medium (5-6) and high (7-10).

The MHS Immunocompromised Registry captures all members with at least one of the following treatments or diagnoses: organ or bone marrow transplant, immunosuppressive treatment (oncologic or other), advanced kidney disease (advanced chronic kidney disease, dialysis, Nephrotic Syndrome), Asplenia or any records of immunosuppression drug purchase.

COVID-19 related symptoms were described as present at first or second PCR test result. Among individuals with a COVID-19 related hospital record, those with a hospitalization within 10 days of their first or second positive PCR test were accounted for and described.

### Descriptives

In addition to individual characteristics, the number of individuals (i.e. individual counts) were assessed and displayed in figures. Age distribution was presented, as reinfection counts, by ranges of 10 years. Distribution of the second positive PCR test was displayed per month. Lastly, the number of days between first and second positive PCR test was evaluated and shown by ranges of 50 days (with a minimum of 100 to 149 days).

## Results

A total of 149,735 individuals in MHS had a record of a positive PCR test between March 2020 and January 2021. Among them 154 MHS members had two positive PCR tests at least 100 days apart and were included in this study, amounting to 0.1% proportion of reinfection. In this cohort, 73 individuals (47.4%) had symptoms at both PCR positive events. Members’ characteristics are displayed in Table 1.

**Table 1.**
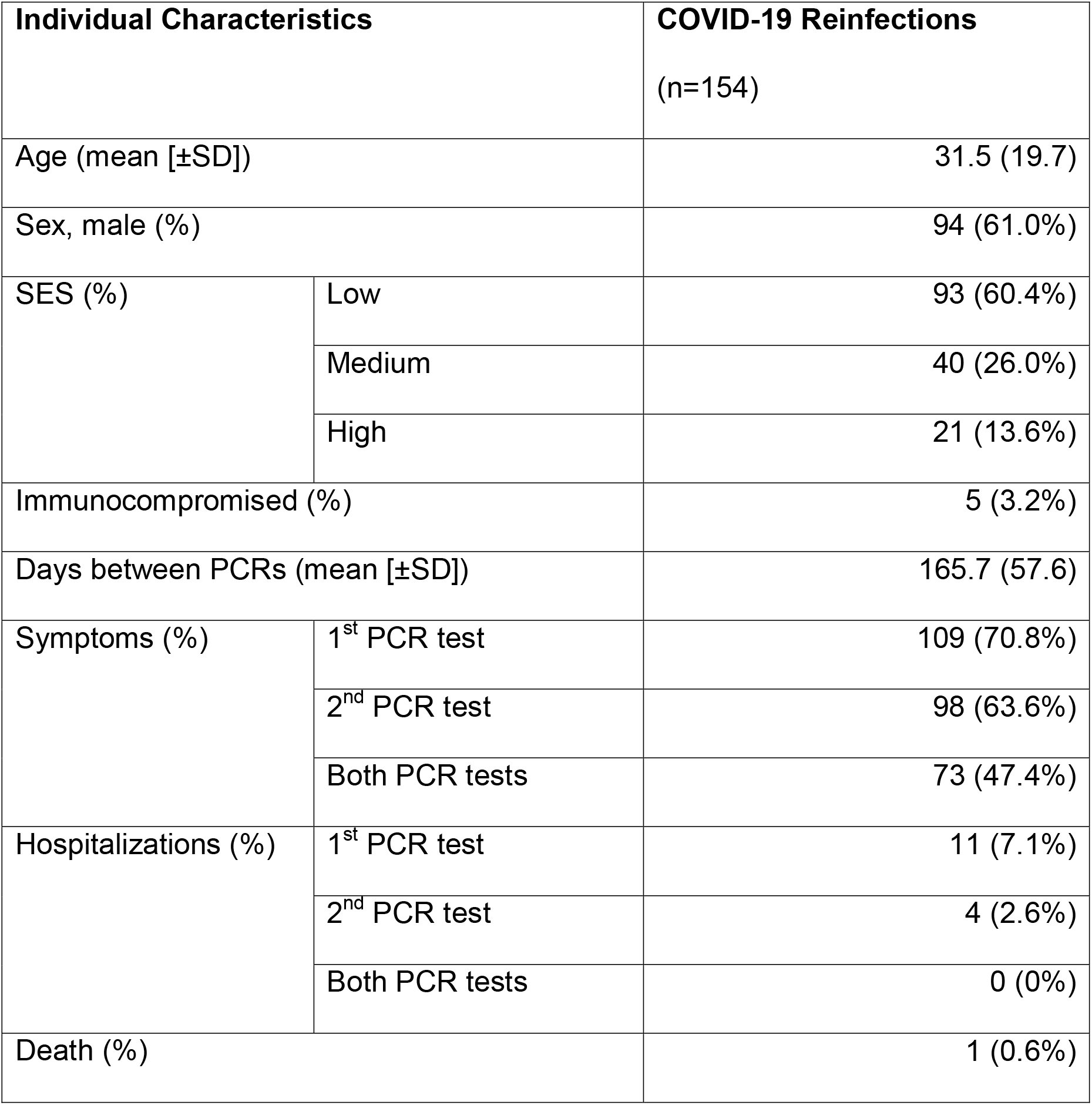
Descriptive results for individuals with COVID-19 reinfections in MHS

Age distribution is presented in Figure 1, where the highest count was among individuals aged 10 to 19 years old. The monthly distribution (Figure 2) displayed that the first reinfection (1 member) occurred in July 2020, where the reinfection counts peak in January 2021 (99 members). The distribution of days between first and second positive PCR (Figure 3) shows that 30 individuals were reinfected more than 200 days following their first positive PCR test.

**Figure 1.**
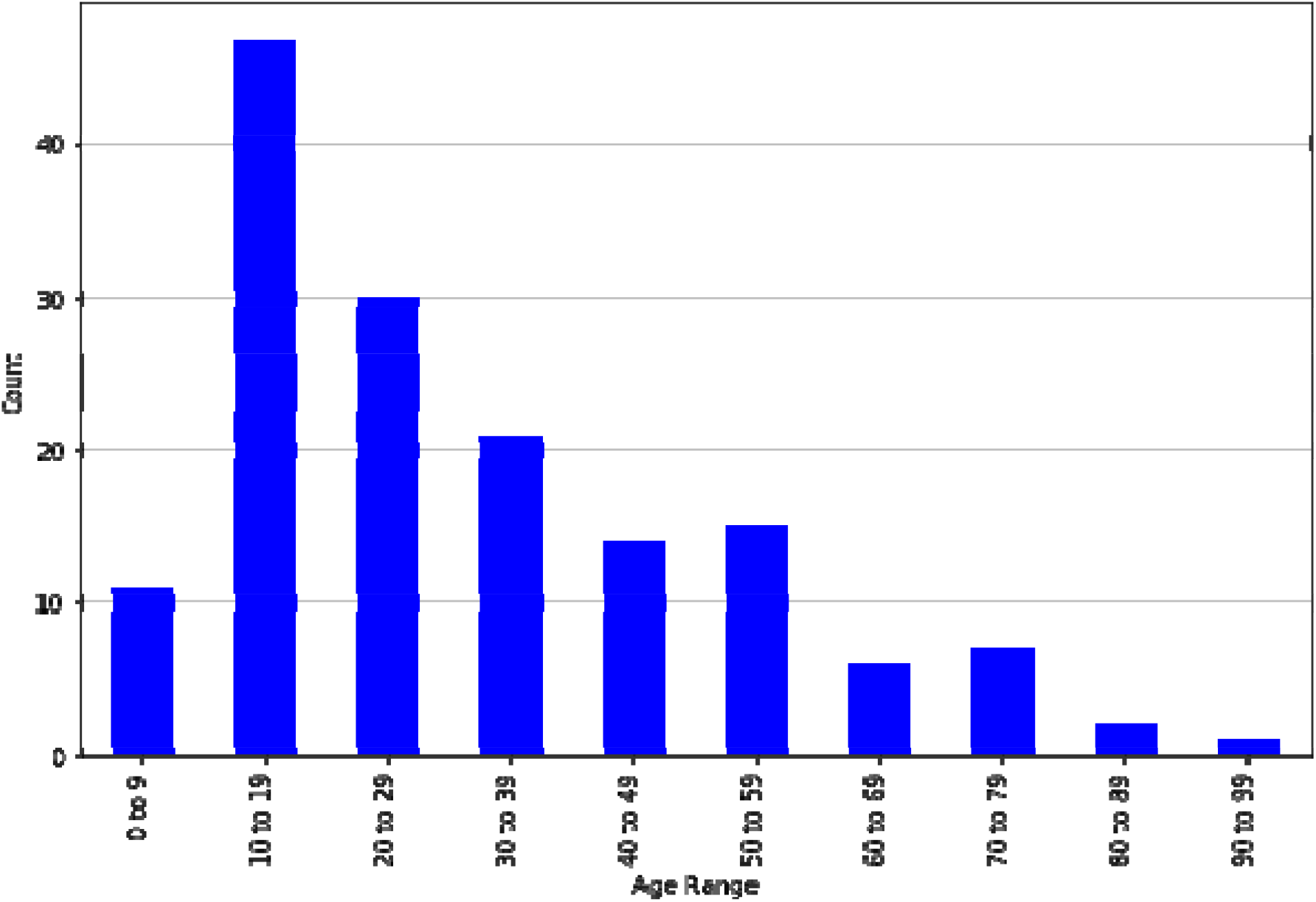
Age distribution among individuals with a COVID-19 reinfection (n=154)

**Figure 2.**
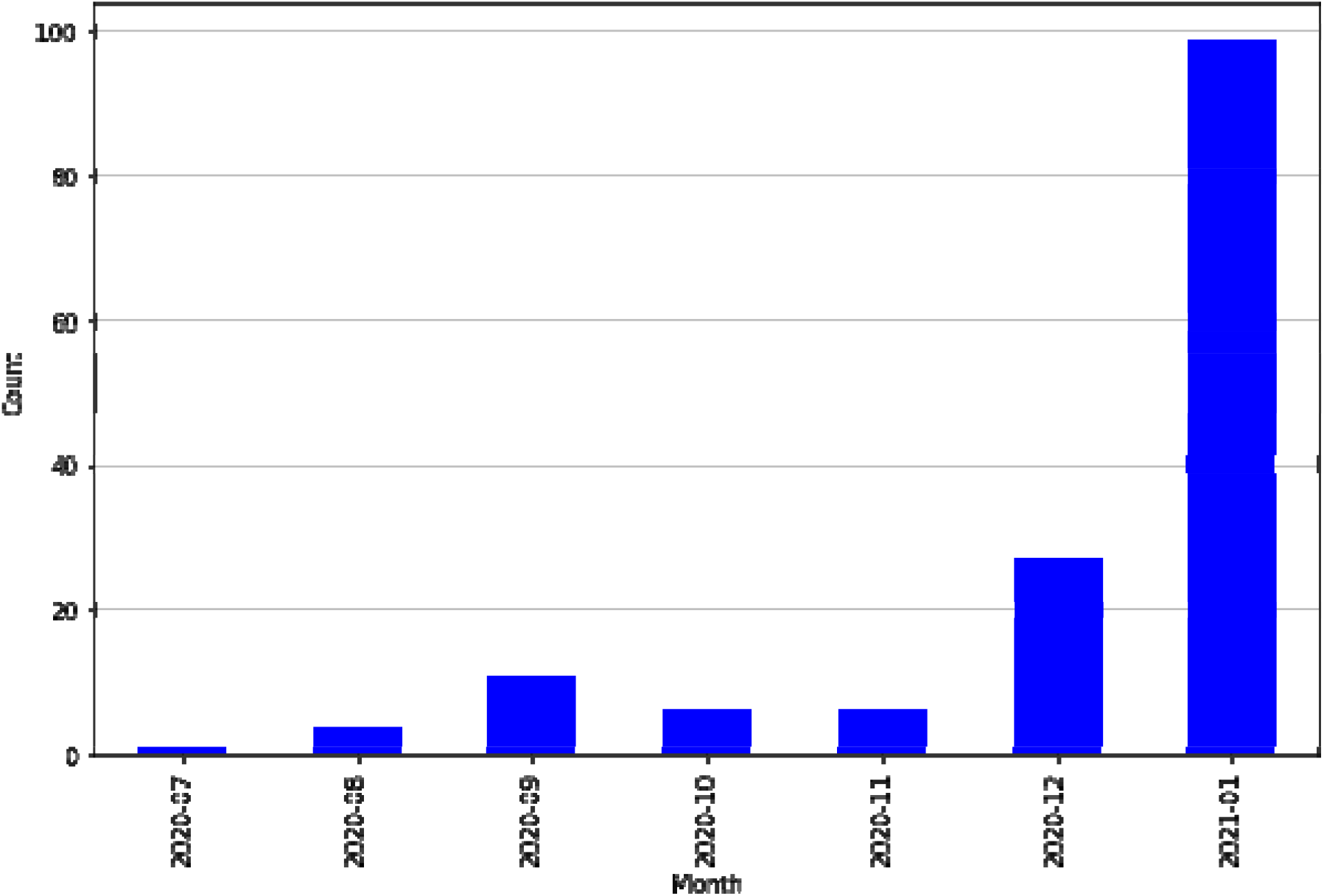
Monthly distribution of second positive PCR test among individuals with a COVID-19 reinfection (n=154)

**Figure 3.**
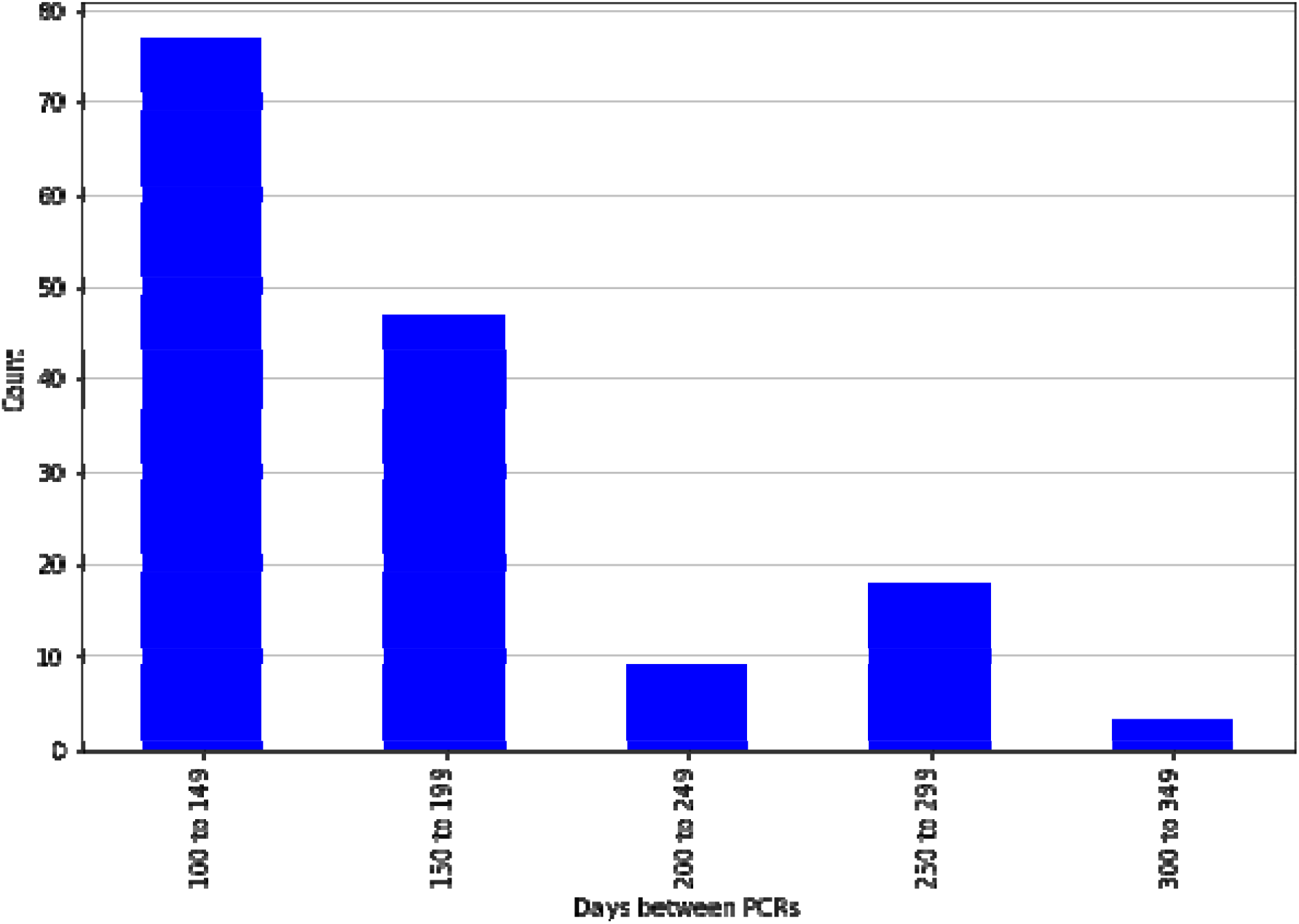
Distribution of days between first and second positive PCRs among individuals with a COVID-19 reinfection (n=154)

## Discussion

A yet unanswered question is the definition of COVID-19 reinfection. While clear cut cases exist, namely two different clinical occasions with two distinct sequenced viruses, relying solely on these will likely cause a massive under-estimation of the reinfection phenomena. Different criteria have been suggested and used in studies, trying to identify true reinfections using easier-to-obtain information and separate those with cases of prolonged viral shedding^[11, 12]^.

This descriptive preliminary report strongly suggests that COVID-19 reinfections have occurred in MHS in Israel. Out of 149,735 individuals with a documented positive PCR test between March 2020 and January 2021, 154 had two positive PCR tests at least 100 days apart, reflecting a reinfection proportion of 1 per 1000. We believe these numbers represent true reinfection incidence in MHS and should be clinically regarded as such.

Admittedly, without genome sequencing, COVID-19 reinfections cannot be conclusively determined; however, we chose a very strict 100-day period and risk factors previously shown to be associated with prolonged viral shedding (especially mechanical ventilation, late hospital admission and immunosuppression) were not prevalent in the population meeting our inclusion criteria ^[13-15]^. In addition, 73 members (47.4%) reported symptoms at two positive PCR tests, suggesting they showed clinical signs of COVID-19 reinfection at both events. The presence of symptoms also reinforces the suggested reinfection among members. We are conducting further research to verify our inclusion criteria.

The peak reinfection counts from January 2021 (Figure 3) should also be addressed. A possible hypothesis for the elevated reinfection counts may refer to the presence of different COVID-19 strains. Multiple genetically distinct variants of SARS-CoV-2 have been documented^[16]^ around the world, as coronavirus mutations were identified in Israel in late November (according to the Israeli Ministry of Health)^[17]^ and become widespread in January^[18-20]^. Correlations between strains and the risk of reinfection should be studied further.

Lastly, the demonstrated age distribution, which suggests higher counts of reinfection among younger individuals, differs from previous analyses that display reinfection rates irrespective of patient age and gender^[3]^. Possible explanations include behavioral factors, namely the lack of social distancing and mask-wearing that may account for an increased exposure and therefore possible increased risk of reinfection.

### Limitations

Although this research displays new information on reinfected COVID-19 individuals in Israel, the study has some limitations. First, the conservative definition of reinfections (i.e. minimum 100 days between positive PCR tests) may have excluded some reinfected members from this study. Furthermore, members in the cohort were not evaluated further than preliminary assessments, mainly counts and proportions. Further research should be performed as more data is collected, followed by statistic modeling and predictive analyses between reinfection groups.

### Conclusions

This study describes real-world data of SARS-CoV-2 reinfection in a large-scale population cohort. Reinfection proportion, albeit small, is not insignificant; as time passes the potential for reinfection increases. Given our strict inclusion criteria, we believe these numbers represent true reinfections in MHS and should be clinically regarded as such.

Health policymakers should acknowledge the possibility of reinfection and reconsider the differential message to recovered population.

## Data Availability

This retrospective study was conducted using data from the Maccabi Healthcare Services (MHS) central computerized database, the second largest state-mandated not-for-profit healthcare provider in Israel, containing more than 2.5 million members (approximately 25% of the population) and is a representative sample of the Israeli population. This fully computerized database captures all information on patient interaction (including demographics, visits, diagnoses, procedures and more). The data is made accessible by MHS Research and Innovation Center under the MHS institutional review board guidelines and approval.

